# Longitudinal Digital Phenotyping of Multiple Sclerosis Severity Using Passively Sensed Behaviors and Ecological Momentary Assessments

**DOI:** 10.1101/2024.11.02.24316647

**Authors:** Zongqi Xia, Prerna Chikersal, Shruthi Venkatesh, Elizabeth Walker, Anind Dey, Mayank Goel

## Abstract

**Background:** Longitudinal tracking of multiple sclerosis (MS) symptoms in an individual’s own environment may improve self-monitoring and clinical management for people with MS (pwMS).

**Objective:** We present a machine learning approach that enables longitudinal monitoring of clinically relevant patient-reported symptoms for pwMS by harnessing passively collected data from sensors in smartphones and fitness trackers.

**Methods:** We divide the collected data into discrete periods for each patient. For each prediction period, we first extract patient-level behavioral features from the current period (action features) and the previous period (context features). Then, we apply a machine learning (ML) approach based on Support Vector Machine with Radial Bias Function Kernel and AdaBoost to predict the presence of depressive symptoms (every two weeks) and high global MS symptom burden, severe fatigue, and poor sleep quality (every four weeks).

**Results:** Between November 16, 2019, and January 24, 2021, 104 pwMS (84.6% women, 93.3% non-Hispanic White, 44.0±11.8 years mean±SD age) from a clinic-based MS cohort completed 12-weeks of data collection, including a subset of 44 pwMS (88.6% women, 95.5% non-Hispanic White, 45.7±11.2 years) who completed 24-weeks of data collection. In total, we collected approximately 12,500 days of passive sensor and behavioral health data from the participants. Among the best-performing models with the least sensor data requirement, ML algorithm predicts depressive symptoms with an accuracy of 80.6% (35.5% improvement over baseline; F1-score: 0.76), high global MS symptom burden with an accuracy of 77.3% (51.3% improvement over baseline; F1-score: 0.77), severe fatigue with an accuracy of 73.8% (45.0% improvement over baseline; F1-score: 0.74), and poor sleep quality with an accuracy of 72.0% (28.1% improvement over baseline; F1-score: 0.70). Further, sensor data were largely sufficient for predicting symptom severity, while the prediction of depressive symptoms benefited from minimal active patient input in the form of response to two brief questions on the day before the prediction point.

**Conclusions:** Our digital phenotyping approach using passive sensors on smartphones and fitness trackers may help patients with real-world, continuous, self-monitoring of common symptoms in their own environment and assist clinicians with better triage of patient needs for timely interventions in MS (and potentially other chronic neurological disorders).

## INTRODUCTION

Multiple Sclerosis (MS) is a leading cause of chronic neurological disability, affecting around 2.8 million people worldwide and over 700,000 people in the United States while causing high health and socioeconomic burdens [1–3]. People with MS (pwMS) may experience a variety of neurological symptoms involving the cognitive, motor, sensory, vision, bowel or bladder domain as well as symptoms of depression, fatigue, and sleep disturbance in their daily lives [4]. Comprehensive MS care involves timely symptom management, but clinician’s awareness of symptoms often lags behind patient experience. Frequent symptom monitoring could improve clinical care and quality of life. However, active engagement with frequent longitudinal symptom monitoring is impractical for patients or clinicians. Given the pervasiveness of MS-related symptoms, symptom monitoring in the patient’s own environment coupled with effective prediction of symptom severity could facilitate triage for timely clinical intervention and reduce the delay in symptom management before worsening.

The digital phenotyping framework uses passively collected data from personal digital devices (*e.g.*, smartphones, fitness trackers) to quantify human behavior moment-by-moment in situ and predict individual health outcomes [5]. Previous works using passively sensed smartphone and wearable data to predict MS outcomes explored the feasibility of passive data collection and the preliminary association between sensed behaviors and standard rater-assessed clinical outcomes [6–14]. However, little is known regarding the clinical applicability of continuous longitudinal digital phenotyping to predict the severity of clinically relevant *patient-reported symptoms* in pwMS. Here, we propose a machine learning approach that harnesses continuously and passively collected data from patients’ digital devices to predict short-term future symptoms. Specifically, we prioritize common MS neurological symptoms as well as symptoms of depression, fatigue, and sleep disturbance that collectively worsen the quality of life.

The primary study goal is to test the feasibility of low-cost, continuous, and longitudinal symptom tracking in a patient’s own environment with minimal active patient engagement. Secondarily, we examined whether machine learning model performance based on passively collected sensor data would improve when (1) using behavioral features from the previous period (context features) to help the models contextualize the patient’s current behaviors in addition to behavioral features from the current period (action features), and (2) incorporating minimal active patient input via response to short surveys called Ecological Momentary Assessments (EMAs). These aspects of the study design in digital phenotyping of clinically relevant patient-reported symptoms differentiate from prior studies. Our approach may also inform the real-world application of long-term, continuous symptom tracking and real-world clinical prediction in chronic neurological conditions beyond MS.

## METHODS

### Participants, Study Period, and Ethics Approval

The study included adults 18 years or older with a neurologist-confirmed MS diagnosis who owned a smartphone (Android or iOS) and enrolled in the Prospective Investigation of Multiple Sclerosis in the Three Rivers Region (PROMOTE) study, a clinic-based MS natural history cohort at the University of Pittsburgh [15–21]. The institutional review boards of the University of Pittsburgh (STUDY19080007) and Carnegie Mellon University (STUDY2019-00000037) approved the study. All participants provided written informed consent. Between November 16, 2019, and January 24, 201, 104 participants completed the data collection for a pre-defined period of 12 weeks, while 44 (out of the 104) participants extended data collection for an additional 12 weeks to complete 24 weeks of data collection. None of the participants experienced acute relapse during the study period. To protect confidentiality, we removed identifiable information (*e.g.*, names, contact information) from sensor and questionnaire data before analysis.

### Overview of the Digital Phenotyping Approach

To briefly summarize the overall approach, we used passively and continuously collected data from participants’ own digital devices, including three *smartphone sensors* (calls, locations, screen usage) and three *fitness tracker sensors* (heart rate, sleep, steps), to predict short-term future patient-reported symptoms of MS-related global neurological symptom burden, depression, fatigue, and sleep quality. To assess the added predictive utility of Ecological Momentary Assessments (*EMAs*), which were brief surveys for “repeated sampling of subjects’ current behaviors and experiences in real-time in subjects’ natural environments” [22, 23], we administered EMAs three times per day through a mobile application asking two multi-choice questions that took less than 15 seconds on average to respond. To capture the real-world fluctuation in symptom severity, we divided each participant’s collected data into discrete consecutive periods (*e.g.,* 2 or 4 weeks) for rolling predictions of patient-reported symptoms. We used patient’s response to validated symptom questionnaires during the same period as the ground truth of symptom severity. We computed features from the sensor and EMA data and classified features as action versus context based on the temporal relationship between features and patient-reported symptom severity at each period. **Action features** captured a person’s activity and behaviors during the period immediately preceding the *next* point of symptom severity prediction. **Context features** captured a person’s activity and behaviors during the period immediately preceding the *previous* prediction point, *i.e.,* the context of a participant’s action features. We then used (1) action features or (2) action and context features to predict the symptom severity.

### Sensor and EMA Data Collection

At enrollment, the study team helped each participant install a custom-built mobile application on their smartphone. In parallel, the study team provided each participant a Fitbit Inspire HR device to wear. Participants kept the Fitbit after study completion. We asked participants to always carry their smartphones, wear fitness trackers, and keep their devices charged.

The mobile application based on the AWARE framework [24] provided the backend and network infrastructure for unobtrusively collecting call logs (*e.g.*, incoming, outgoing, and missed calls), locations, and screen usage (*i.e.,* when the screen status changed to on or off and locked or unlocked) of the smartphone sensors. The fitness tracker sensors captured heart rate, sleep status (*e.g.,* asleep, awake, restless, or unknown), and the number of steps. Data from AWARE were deidentified and automatically transferred over WiFi to a study server at regular intervals. Data from the Fitbit were retrieved using the Fitbit Application Programming Interface at the end of each participant’s data collection.

Calls and screen usage were event-based sensor streams, whereas location, heart rate, sleep, and steps were time series sensor streams. We sampled location coordinates at 1 sample per 10 minutes and heart rate, sleep, and steps at 1 sample per minute.

Throughout the study duration, the mobile application alerted and directed participants three times a day to complete a brief EMA survey within the application. EMA surveys took less than 15 seconds to complete on average. The two recurring questions were: (1) “How depressed do you feel?” and (2) “How tired do you feel?”. Participants responded to each EMA question using a Likert scale from 0 to 4, with 0 indicating the least and 4 indicating the most depressed/tired feeling. The EMA responses were transmitted to the study server.

### Questionnaire Deployment for Assessing Symptom Severity

Participants completed online questionnaires using the secure, web-based Research Electronic Data Capture (REDCap) system [25, 26]. To assess the severity of clinically relevant symptoms, we used standardized patient-reported outcome questionnaires validated in pwMS. To harmonize the periods across participants, all participants completed a baseline questionnaire assessing demographics and clinical profiles on the Saturday following enrollment. Beyond the baseline, participants completed additional questionnaires at regular intervals (*e.g.,* every 2 or 4 weeks from the first Saturday) as appropriate for assessing each standard patient-reported symptom type throughout the data collection period.

#### Depressive Symptom

To measure the severity of depression symptoms, participants completed the Patient Health Questionnaire (PHQ-9) *once every two weeks* [27]. The PHQ-9 asked for symptoms in the preceding two weeks, whereas the other questionnaires in this study asked for symptoms in the preceding four weeks. PHQ-9 scores ranged from 0 to 3, with higher scores indicating more severe depressive symptoms.

#### Global MS Neurological Symptom Burden

To measure the severity of the global MS-related neurological symptom burden, participants completed the Multiple Sclerosis Rating Scale-Revised (MSRS-R) *once every four weeks* [28]. MSRS-R assessed eight neurological domains (*i.e.,* walking, upper limb function, vision, speech, swallowing, cognition, sensory, bladder, and bowel function). Each domain could score from 0 to 4, with 0 indicating the absence of symptoms and 4 indicating the greatest symptom severity. The total score (0 to 32) indicates the global MS-related neurological symptom burden.

#### Fatigue

To measure the severity of fatigue, participants completed the 5-item version of the Modified Fatigue Impact Scale (MFIS-5) *once every four weeks* [29]. MFIS-5 assessed the impact of fatigue on cognitive, physical, and psychosocial function. Each item in MFIS-5 could score from 0 (never) to 4 (almost always) on a five-point Likert scale, with higher scores indicating more severe fatigue.

#### Sleep Quality

To measure the severity of sleep disturbances, participants completed the Pittsburgh Sleep Quality Index (PSQI) *once every four weeks* [30]. The 19 items of PSQI generated 7 component scores (each on a 0-3 scale) and one composite score (0 to 21), with higher scores indicating poorer sleep quality.

Binary indicators of symptom severity likely have more practical real-world clinical utility in assisting patient self-monitoring and facilitating clinician triage for symptom intervention. For each symptom type, we dichotomized the score to the respective standardized questionnaire using specific thresholds to classify symptom severity. For global MS neurological symptom burden, we dichotomized MSRS-R scores as ≥6.4 (higher burden) versus <6.4 (lower burden). For depressive symptoms, we dichotomized PHQ-9 scores as ≥5 (presence of depressive symptoms) versus <5 (absence of depressive symptoms). For fatigue, we dichotomized MSIF-5 scores as ≥8 (greater fatigue) versus <8 (lower fatigue). For sleep quality, we dichotomized PSQI scores as ≥9 (poorer sleep quality) and <9 (better sleep quality). For depressive symptoms and sleep quality, the binary thresholds were based on previous consensus [27, 31]. For global MS neurological symptom burden and fatigue, we calculated the respective median scores in the entire dataset. We used the median scores as the thresholds, given the lack of consensus from the literature. Thus, throughout the data collection, each participant has a consecutive series of binary symptom severity status (*i.e.,* every two weeks for depressive symptoms and every four weeks for global MS symptom burden, fatigue and sleep quality).

### Sensor and EMA Data Processing and Machine Learning

Briefly, the data processing and analysis pipeline required the following steps (Figure 1). First, we extracted features from sensor and EMA data to generate action and context features. Second, we improved data quality by handling missing features. Finally, we implemented a machine learning pipeline to predict the severity of each patient-reported symptom on a rolling basis (*i.e.,* every two weeks for depressive symptoms and every four weeks for global MS neurological symptom burden, fatigue and sleep quality) using action features or action + context features in the following iterations: (1) 1-sensor models, each containing features from one out of the six sensor types; (2) the best combination of the 1-sensor models; (3) the best combination of 1-sensor models plus EMA.

**Figure 1.**
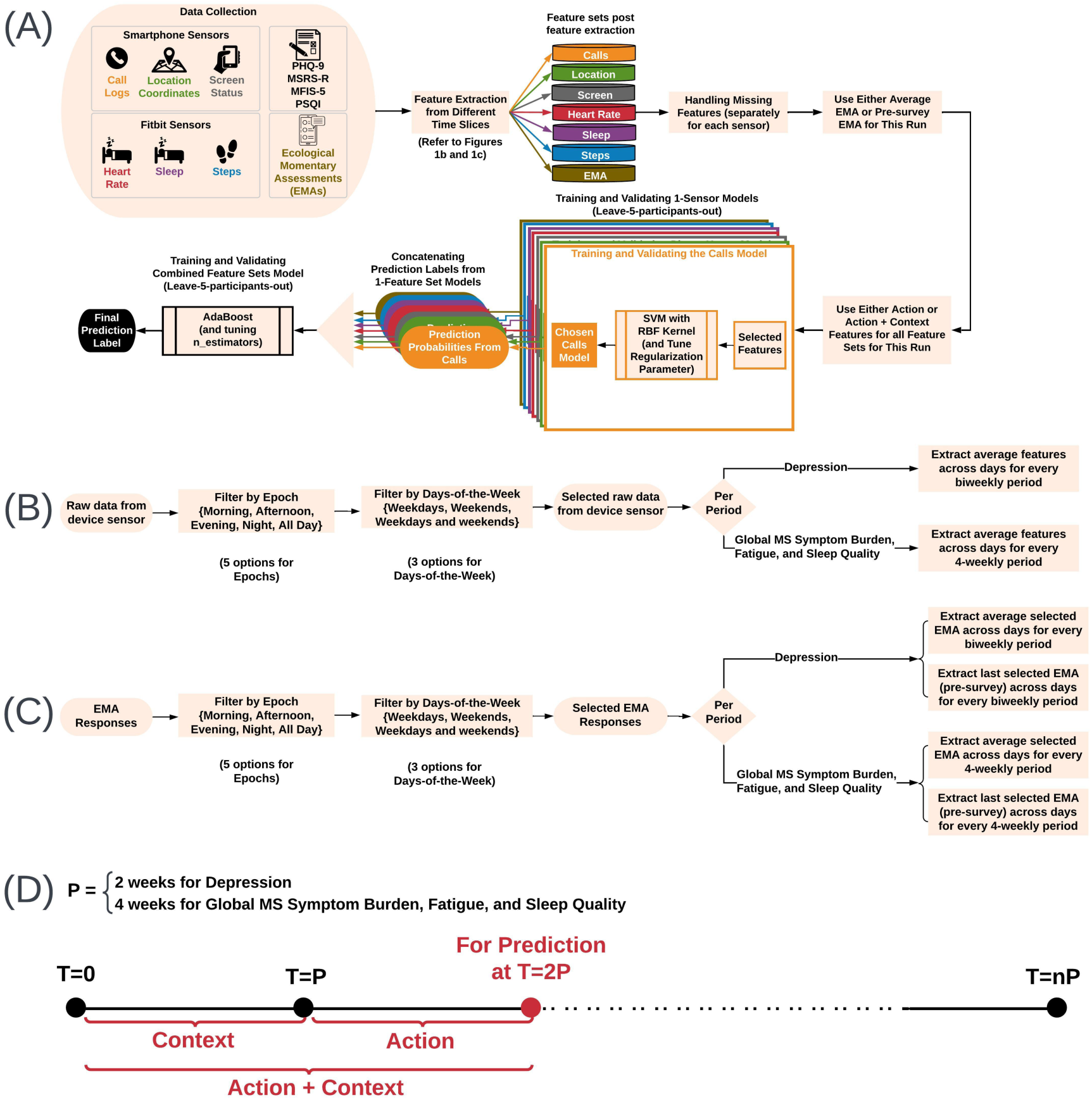
Data Processing and Analysis Pipeline. (A) The pipeline for predicting depressive symptoms (PHQ-9) every 2 weeks, and global MS neurological symptom burden (MSRS-R), fatigue (MFIS-5) and sleep quality (PSQI) every 4 weeks, used passively collected sensor data from smartphones and fitness trackers as well as EMAs. We ran the pipeline for two types of EMA features (average and pre-survey EMAs) and two types of feature matrices (action and action + context). (B) For each sensor, every feature was extracted from 15 temporal slices over 2 or 4-week periods. First, raw data from the device sensor were preprocessed and filtered by time-of-the-day and days-of-the-week. Features were then extracted from the selected raw data. (C) For EMA, we used a similar approach (as for processing sensor data) to calculate the average EMA and pre-survey EMA. (D) Action features were features from the period immediately preceding the prediction point, whereas context features were from the period preceding the “action period”.

#### Feature Extraction and Engineering

*Overview:* From the smartphone and fitness tracker sensors, we computed six types of features from different sensors (*i.e.,* Calls, Heart Rate, Location, Screen, Sleep, and Steps), given their known potential to inform behaviors relevant to symptoms of depression [32–37], fatigue [10], poor sleep quality [38, 39] and crucial MS neurological symptoms such as decreased mobility [13]. The “Calls” features captured communication patterns. The “Heart Rate” and “Steps” features captured the extent of physical activities. The “Location” features captured mobility patterns. The “Screen” features potentially captured the ability for concentration [40, 41] and the extent of sedentary behavior [42] with caveats for pwMS and people with other chronic neurological disorders who may experience impairment with upper limb or fine motor functions. The “Sleep” features captured sleep duration and patterns, from which we could infer sleep disturbance (*e.g.*, insomnia or hypersomnia) [43]. The **Supplementary Materials Section A.1** provided details of sensor feature extraction and engineering. For sensor features over time periods (*e.g.,* every 2-week or 4-week period) (Figure 1B), we calculated the daily average value of each sensor feature. Given the diversity of behaviors with ephemeral and sustained changes in pwMS, it is crucial to initialize the model with a large feature set. While these features captured individual or overlapping behaviors, the feature selection stage of our machine learning pipeline removed redundant features.

For EMA responses during the same time periods (*e.g.,* every 2-week or 4-week period) (Figure 1C), we obtained two types of EMA features. The “Average EMA” was the daily average value of each EMA question response during a given period. The “Pre-survey EMA” represented the value of the last response to each EMA question on the day prior to the administration of the questionnaire for assessing the patient-reported symptom during each period.

*Temporal Slicing:* The temporal slicing approach extracted sensor features from different time segments (Figure 1B and 1C). From prior research, temporal slicing better defined the relationship between a sensor feature and depression severity, for example [44, 45]. Here, we collected all available data during each specific epoch or time segment of the day (all day, night [00:00-06:00 hours], morning [06:00-12:00 hours], afternoon [12:00-18:00 hours], evening [18:00-00:00 hours]) and on specific days of the week (all days of the week, weekdays only [Monday-Friday], weekends only [Saturday-Sunday]) to achieve 15 data streams or **temporal slices**. For sensor or EMA features in each of the 15 temporal slices, we first computed daily features (of the temporal slice) and averaged daily features over either 2- or 4-week periods for prediction (*i.e.,* every two weeks to predict depressive symptoms and every four weeks to predict global MS neurological symptom burden, fatigue, and sleep quality). We concatenated the features from 15 temporal slices to derive the final feature matrix.

*Feature Matrix:* After feature extraction, we created a feature matrix for each of the six sensors (calls, locations, screen usage, heart rate, sleep, steps) and each of the two EMA types (average and pre-survey EMA), containing features for the 15 temporal slices in consecutive 2 or 4-week periods during each participant’s study follow-up. The “action” feature matrix captured each participant’s actions during the *current* (2 or 4-week) period, at the end of which period we predicted the patient-reported symptom severity as the outcome. For each participant, we concatenate features from the *previous* (2 or 4-week) period, which captured the context for the current actions, with the “action” feature matrix to obtain the “action + context” feature matrix. Thus, to predict the outcome at the end of the i^th^ period at time T = iP where P = 2 weeks or four weeks, the action feature matrix comprised features from time (i-1)P and time iP, whereas the action + context feature matrix comprised features from time (i-2)P and time iP (Figure 1D).

#### Handling Missing Data

Missing sensor data could occasionally occur due to several reasons. **Supplementary Materials Section A.2** described the detailed approach for handling missing data.

#### Machine Learning (ML) Pipeline Using Action and Context Behavioral Features

We built machine learning models using Support Vector Machines (SVM) with Radial Bias Function (RBF) Kernels and validated our models using *leave-5-participants-out cross-validation* to mitigate over-fitting. As an overview, the pipeline involved six steps. First, in the *Generating Feature Sets* step, we created model configurations that enabled assessment of the utility of EMA features and contextual feature information. Second, we performed *Training and Validating 1-Sensor and EMA-only Models* step for each of the six sensor feature types (Calls, Heart Rate, Location, Screen, Sleep, and Steps) and either EMA feature type (average or pre-survey EMA features). Third, during the *Obtaining Predictions from Combinations of Sensors* step, we combined detection probabilities from 1-sensor models to identify the best performing combined sensor model. Fourth, during *Obtaining Predictions from Combinations of Sensors and EMA* step, we combined detection probabilities from 1-sensor models and an EMA-only model to identify the best performing final model. Fifth, we performed the *Classifying Different Outcomes* step by running the pipeline for each outcome. Finally, we performed a comparison of ML models using *Bootstrapping Predictions*.

*Generating Feature Sets:* We generated features for the different model configurations to assess the utility of EMA features and contextual feature information. For *EMA,* we used (1) no EMA information, (2) only pre-survey EMA, or (3) average EMA values. For *Context*, we either used (1) only Action or (2) Action+Context. In total, there were six configurations based on these features.

*Training and Validating 1-Sensor and EMA-only Models:* For each sensor and EMA feature matrix, we built a model of the selected features from the given sensor or EMA type to predict an outcome (Figure 2). We trained models using an SVM classifier with RBF Kernel (SVM-RBF). We used leave-5-participants-out cross-validation to choose the regularization parameter for SVM-RBF. The folds were split in a stratified manner, and classes were balanced in the SVM-RBF to ensure that positive and negative classes of the binary outcomes were adequately represented. We chose the model with the best F1-score for a given outcome, which provided the prediction probabilities for the outcome. The process for one outcome was independent of the other outcomes.

**Figure 2.**
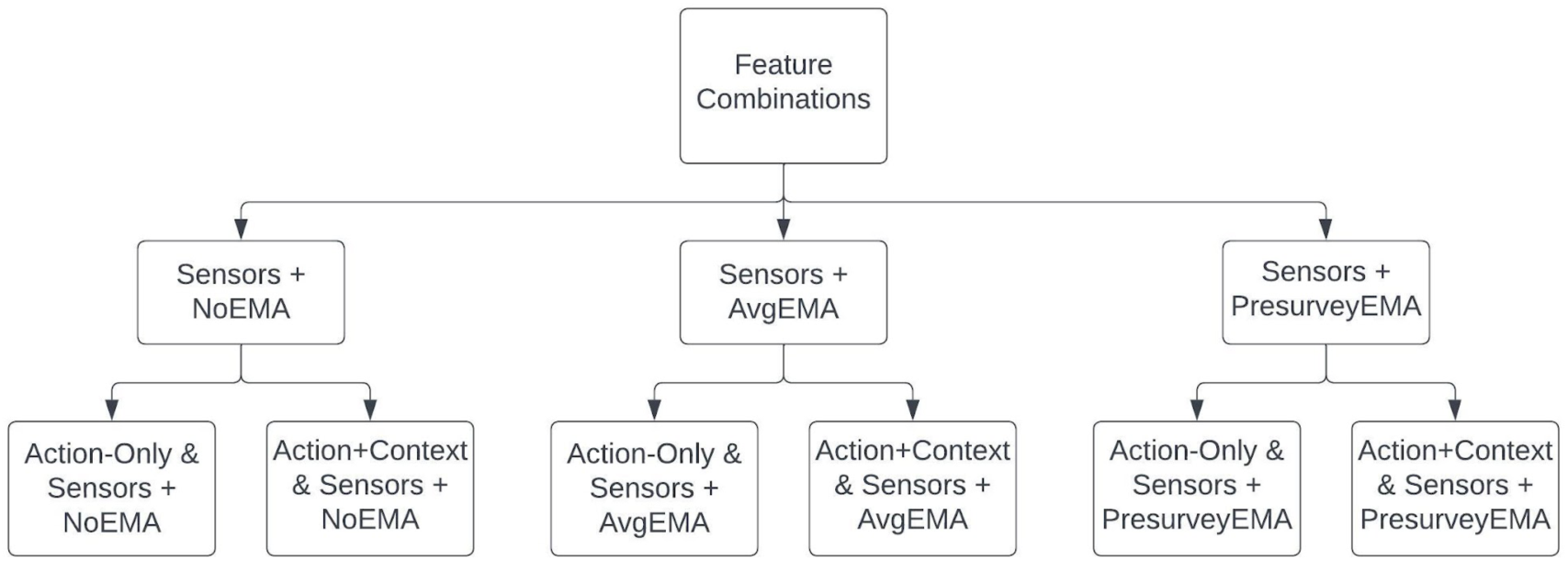
Feature combinations from sensors and EMAs.

*Obtaining Predictions from Combinations of Sensors:* We concatenated prediction probabilities from all six 1-sensor models into a single feature vector and entered as input into an ensemble classifier, *i.e.*, AdaBoost with Decision Tree Classifier as a base estimator, which generated the final prediction for each outcome. For all outcomes, only the prediction probabilities of the positive label “1” were concatenated. The positive labels were the “presence of depressive symptoms” for depression, “high burden” for global MS neurological symptom burden, “severe fatigue” for fatigue, and “poor sleep quality” for sleep quality. We tuned the “n_estimators” (*i.e.,* the maximum number of estimators at which boosting was terminated) parameter during leave-5-participants-out cross-validation to achieve the best-performing combined model.

To analyze the contribution of each sensor combination, we implemented a feature ablation analysis by generating detection results for all possible combinations of 1-sensor models. For six 1-sensor models, there were 57 combinations of feature sets, as the total combinations = combinations with two sensors + … + combinations with six sensors:

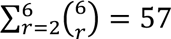

*Obtaining Predictions from Combinations of Sensors and EMA-only Models:* We concatenated prediction probabilities from all six 1-sensor models and one EMA-only model into a single feature vector and entered as input into an ensemble classifier using the same method for sensors (as described above) to train this combined classifier.

To analyze the utility of each sensor and EMA combination, we implemented a feature ablation analysis by generating detection results for all possible combinations of 1-sensor models and the EMA model. For six 1-sensor models and one EMA model, there were 120 combinations of feature sets, as the total combinations = combinations with two sensors or 1 sensor and EMA + … + combinations with six sensors and/or EMA:

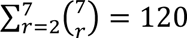

*Classifying Different Outcomes:* We ran the following pipeline independently for each of the four patient-reported symptoms as the outcomes, first using action-only features and then using action + context features:

1. Training and validating six 1-sensor models without EMA and 57 combined models.
2. Training and validating six 1-sensor models plus average EMA and 120 combined models.
3. Training and validating six 1-sensor models plus pre-survey EMA and 120 combined models.

Each patient had multiple “samples” (*i.e.,* prediction periods) over the study duration. For each patient-reported symptom, we trained six final models based on whether the model included Action versus Action + Context features or whether the model contained no EMA, average EMA, or pre-survey EMA. Here, the “positive” label refers to the outcome of interest (*e.g.,* presence of depressive symptoms, presence of high global MS neurological symptom burden, presence of severe fatigue, presence of poor sleep quality). For each final model of a given outcome, we reported the model performance of the best combination of sensors and/or EMA. We also reported the performance of baseline models (*i.e.*, a simple majority classifier whereby every point was assigned to whichever was in the majority in the training set) as well as models containing all six sensors or all six sensors plus one EMA type.

*Comparing ML Models by Bootstrapping Predictions:* For model performance metrics, we assessed accuracy and F1-score. **Accuracy** is the percentage of samples for which the model correctly predicted the outcome label. **F1-score** measures the harmonic mean of precision and recall. Precision is the positive predictive value, *i.e.,* the number of true positive labels divided by the number of all positive labels (true positive + false positive). Recall is sensitivity, *i.e.,* the number of true positive labels divided by the number of all samples that should have the positive labels (true positive + false negative). For each patient-reported symptom, we compared the bootstrapped accuracy and F1 scores among the six final models in a pairwise manner (30 comparisons). Specifically, we computed the 95% confidence intervals of differences in their bootstrapped accuracy and F1-score. We performed hierarchical bootstrapping by randomly sampling {participant ID, prediction week} with replacement over 10000 iterations. In each iteration, we took samples with the same {participant ID, prediction week} across the two models being compared and computed the difference in accuracy and difference in F1-score, respectively. After computing all iterations, we generated the 95% confidence intervals of the difference in accuracy and difference in F1-score (two-tailed alpha = 0.05). If one of the models in a pair is not statistically better than the other, we consider the model requiring the least amount of sensor and/or EMA data to be “better.”

## RESULTS

### Patient Profile

The study included 104 pwMS who completed at least 12-weeks of data collection between November 2019 and January 2021. The subset of the participants who completed 24-weeks of data collection shared similar characteristics as the study cohort, which was largely representative of the larger clinic-based MS population (Table 1).

**Table 1.**
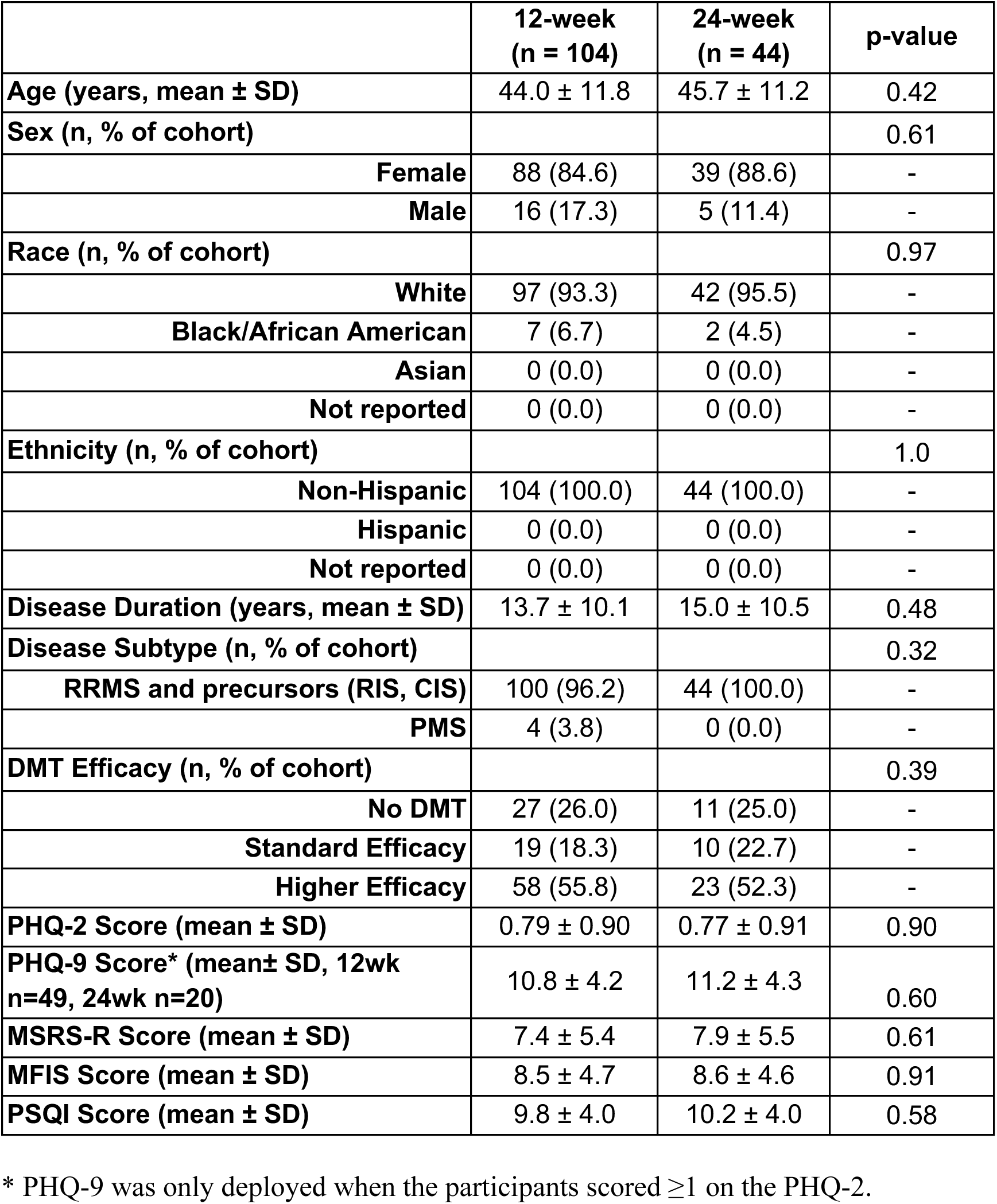
Patient Characteristics.

### Predicting Outcomes Using Action and Context Features from Sensor and EMAs

We report the accuracy and F1-score of the machine learning pipeline for predicting each type of patient-reported symptom using the best-performing sensor and/or EMA combinations (*i.e.*, the set of sensors and/or average or pre-survey EMA) for models trained on Action-only features and Action + Context features (Figure 3). Separately, we report the performance of *individual* 1-sensor, average EMA, and pre-survey EMA models (Supplementary Table S1) as well as models combining all 6 sensors, 6 sensors + average EMA, or 6 sensors + pre-survey EMA (Supplementary Table S2). Finally, we indicate the best combination of sensors and/or EMA, selected for each model type (Supplementary Table S3, corresponding to Figure 3).

**Figure 3.**
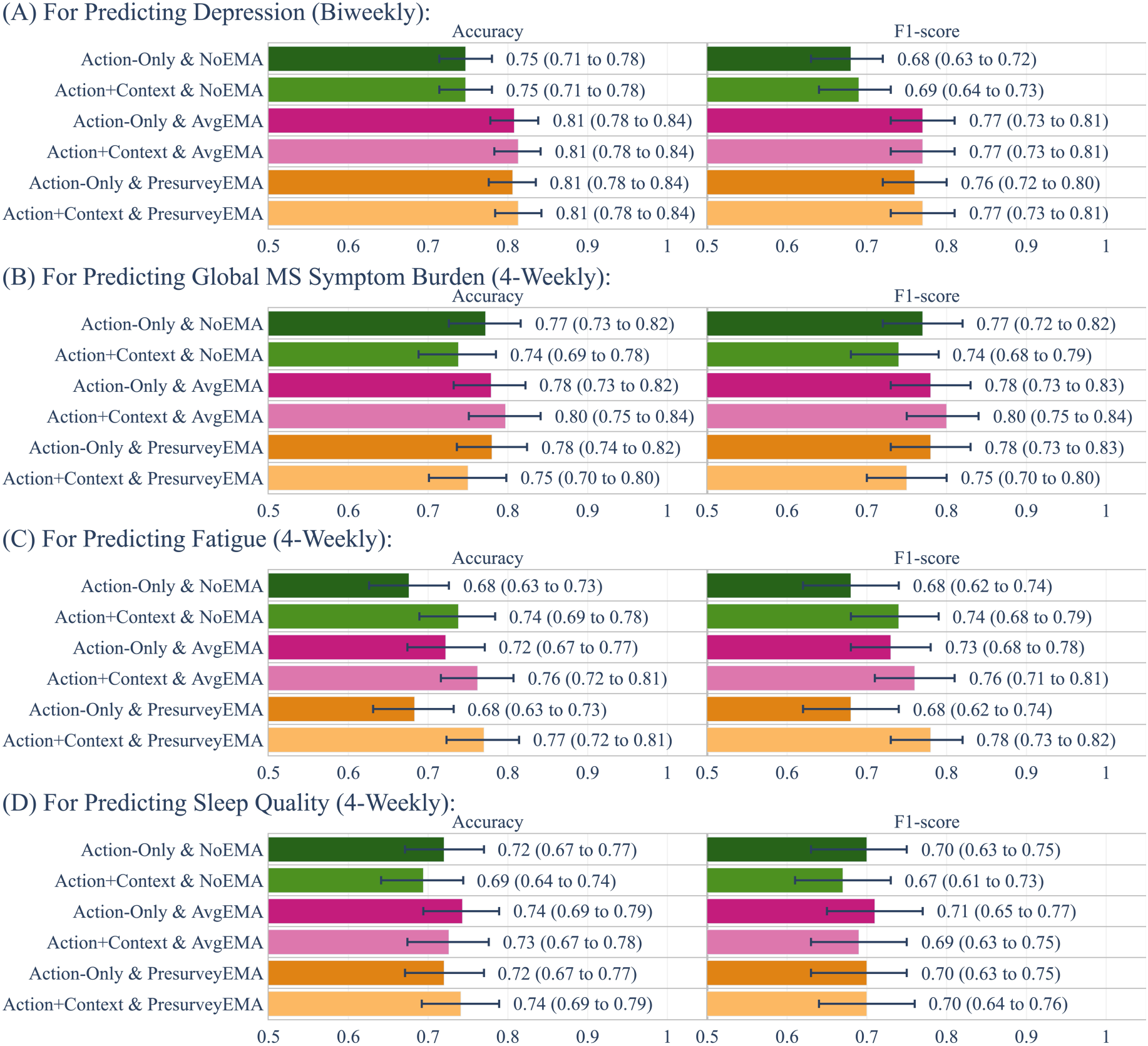
Performance of the machine learning pipeline. We use the best sensor or sensor+EMA combinations for predicting the four patient-reported symptoms in pwMS: (A) depressive symptom, (B) global MS neurological symptom burden, (C) fatigue, and (D) sleep quality. “Action-Only & NoEMA” is the best model that combines predictions of 1-sensor models trained on action-only features. “Action+Context & NoEMA” is the best model that combines predictions of 1-sensor models trained on action and context features. “Action-Only & AvgEMA” is the best model that combines predictions of 1-sensor models and the average EMA model trained on action-only features. “Action+Context & AvgEMA” is the best model that combines predictions of 1-sensor models and the average EMA model trained on action and context features. “Action-Only & PresurveyEMA” is the best model that combines predictions of 1-sensor models and the pre-survey EMA model trained on action-only features. “Action+Context & PresurveyEMA” is the best model that combines predictions of 1-sensor models and the pre-survey EMA model trained on action and context features. We use bootstrapping to report average Accuracy (X 0.01) and F1-score and the corresponding 95% confidence intervals (alpha=0.05) for each of these models.

#### Depressive Symptom

For predicting the presence of depressive symptoms (versus the absence of depressive symptoms) every 2 weeks, the *baseline* model (simple majority classifier) had an accuracy of 59.5%. The model containing *all 6 sensors and no EMA* had an accuracy of 74.7% with action-only features (25.5% relative improvement over the baseline), and an accuracy of 72.2% with action + context features (21.3% relative improvement over the baseline) (Supplementary Table S2). The model containing the *best combination of sensors and no EMA* had an accuracy of 74.7% with action-only features (25.5% relative improvement over the baseline; Best combination: calls, heart rate, location, screen, sleep, and steps), and an accuracy of 74.7% with action + context features (25.5% relative improvement over the baseline; Best combination: calls, heart rate, location, screen, and sleep) (Figure 3). The model containing the *best combination of sensors and average EMA* had an accuracy of 80.8% with action-only features (35.8% relative improvement over the baseline; Best combination: heart rate, sleep, steps, and average EMA), and an accuracy of 81.3% with action + context features (36.6% relative improvement over the baseline; Best combination: calls, heart rate, location, sleep, and average EMA). The model containing the *best combination of sensors and pre-survey EMA* had an accuracy of 80.6% with action-only features (35.5% relative improvement over the baseline; Best combination: heart rate, steps, and pre-survey EMA) and an accuracy of 81.4% with action + context features (36.8% relative improvement over the baseline; Best combination: heart rate, location, screen, and pre-survey EMA).

When comparing the model performance in a pairwise manner (Figure 3), Action+Context & PresurveyEMA had the highest bootstrapped average Accuracy of 81.4% and the highest average F1-score of 0.77. This model significantly outperformed both NoEMA models: Action-Only & NoEMA (absolute increase of 6.7% in accuracy and 0.09 in F1-score), Action+Context & NoEMA (absolute increase of 6.6% in accuracy and 0.1 in F1). Likewise, Action-Only & PresurveyEMA significantly outperformed both NoEMA models: Action-Only & NoEMA (absolute increase of 6.0% in accuracy and 0.09 in F1-score), Action+Context & NoEMA (absolute increase of 6.1% in accuracy and 0.09 in F1-score). Models with average EMA (Action-Only & AvgEMA, Action+Context & AvgEMA) also significantly outperformed both NoEMA models. However, there were no statistically significant differences between Action-Only & PresurveyEMA and Action+Context & PresurveyEMA or between any of the PresurveyEMA models and the AvgEMA models.

Thus, for predicting the presence of depressive symptoms every 2 weeks, the **Action-Only & PresurveyEMA model** generated the best performance (accuracy: 80.6%; F1-score: 0.76) while requiring the least amount of sensor (*e.g.,* heart rate, steps) and EMA data (*e.g.,* pre-survey EMA). Pre-survey EMA was the last EMA response on the day prior to survey completion to assess patient-reported depressive symptoms.

#### Global MS Neurological Symptom Burden

For predicting high global MS neurological symptom burden (vs. low burden) every 4 weeks, the baseline model had an accuracy of 51.1%. The model containing *all 6 sensors and no EMA* had an accuracy of 70.7% with action-only features (38.4% relative improvement over the baseline), and an accuracy of 72.0% with action + context features (40.9% relative improvement over the baseline) (Supplementary Table S2). The model containing the *best combination of sensors and no EMA* had an accuracy of 77.3% with action-only features (51.3% relative improvement over the baseline; Best combination: heart rate, location, sleep, and steps), and an accuracy of 73.8% with action + context features (44.4% relative improvement over the baseline; Best combination: heart rate, location, and sleep) (Figure 3). The model containing the *best combination of sensors and average EMA* had an accuracy of 77.9% with action-only features (52.4% relative improvement over the baseline; Best combination: heart rate, location, sleep, steps, and average EMA), and an accuracy of 79.7% with action + context features (56.0% relative improvement over the baseline; Best combination: calls, heart rate, screen, sleep, and average EMA). The model containing the *best combination of sensors and pre-survey EMA* had an accuracy of 78.0% with action-only features (52.6% relative improvement over the baseline; Best combination: location, sleep, steps, and pre-survey EMA) and an accuracy of 75.1% with action + context features (47.0% relative improvement over the baseline; Best combination: heart rate, location, screen, sleep, and pre-survey EMA).

When comparing the model performance in a pairwise manner (Figure 3), none was significantly better than the most parsimonious Action-Only & NoEMA model. Thus, for predicting high global MS symptom burden every 4 weeks, the **Action-Only & NoEMA** model generated the best performance (accuracy: 77.3%; F1-score: 0.77) while requiring the least amount of sensor data (*i.e.*, heart rate, location, sleep, and steps; trained on action-only features) and importantly no EMA data (*i.e.,* no active participant input).

#### Fatigue

For predicting severe fatigue (vs. mild fatigue) every 4 weeks, the baseline model had an accuracy of 50.9%. The model containing *all 6 sensors and no EMA* had an accuracy of 60.4% with action-only features (18.7% relative improvement over the baseline), and an accuracy of 69.7% with action + context features (36.9% relative improvement over the baseline) (Supplementary Table S2). The model containing the *best combination of sensors and no EMA* had an accuracy of 67.6% with action-only features (32.8% relative improvement over the baseline; Best combination: calls, heart rate, screen, and steps), and 73.8% with action + context features (45.0% relative improvement over the baseline; Best combination: heart rate, screen, and steps) (Figure 3). The model containing the *best combination of sensors and average EMA* had an accuracy of 72.2% with action-only features (41.9% relative improvement over the baseline; Best combination: heart rate, screen, steps, and average EMA), and an accuracy of 76.1% with action + context features (49.5% relative improvement over the baseline; Best combination: heart rate, screen, sleep, steps, and average EMA). The model containing the *best combination of sensors and pre-survey EMA* had an accuracy of 68.3% with action-only features (34.2% relative improvement over the baseline; Best combination: heart rate, screen, steps, and pre-survey EMA), and an accuracy of 77.1% with action + context features (51.5% relative improvement over the baseline; Best combination: calls, heart rate, screen, steps, and pre-survey EMA).

When comparing the model performance in a pairwise manner (Figure 3), none was significantly better than Action+Context & NoEMA. Thus, for predicting severe fatigue every 4 weeks, the **Action+Context & NoEMA** model generated the best performance (accuracy: 73.8%; F1-score: 0.74) while requiring the least amount of sensor data (*i.e.*, heart rate, screen, and steps; trained on action and context features) and importantly no EMA data (*i.e.,* no active participant input).

#### Sleep Quality

For predicting poor sleep quality (vs. better sleep quality) every 4 weeks, the baseline model had an accuracy of 56.2%. The model containing *all 6 sensors and no EMA* had an accuracy of 58.2% with action-only features (3.6% relative improvement over the baseline), and an accuracy of 68.7% with action + context features (22.2% relative improvement over the baseline) (Supplementary Table S2). The model containing the *best combination of sensors and no EMA* had an accuracy of 72.0% with action-only features (28.1% relative improvement over the baseline; Best combination: heart rate, location, sleep, and steps), and an accuracy 69.5% with action + context features (23.7% relative improvement over the baseline; Best combination: calls, heart rate, sleep, and steps) (Figure 3). The model containing the *best combination of sensors and average EMA* had an accuracy of 74.4% with action-only features (32.4% relative improvement over the baseline; Best combination: heart rate, location, screen, sleep, and average EMA), and an accuracy of 72.7% with action + context features (29.4% relative improvement over the baseline. Best combination: heart rate, location, sleep, steps, and average EMA). The model containing the *best combination of sensors and pre-survey EMA* had an accuracy of 72.0% with action-only features (28.1% relative improvement over the baseline. Best combination: heart rate, location, sleep, and steps while Pre-survey EMA was not selected), and an accuracy of 74.0% with action + context features (31.7% relative improvement over the baseline; Best combination: calls, heart rate, sleep, and pre-survey EMA).

When comparing the model performance in a pairwise manner (Figure 3), none was significantly better than the most parsimonious Action-Only & NoEMA model. Thus, for predicting poor sleep quality every 4 weeks, the **Action-Only & NoEMA** model generated the best performance (accuracy: 72.0%; F1-score: 0.70) while requiring the least amount of sensor data (*i.e.*, heart rate, location, sleep, and steps; trained on action-only features) and importantly no EMA data (*i.e.,* no active participant input).

## DISCUSSION

For the primary goal of this study that analyzed ∼12,500 days of passively and continuously collected data from pwMS, we report the feasibility of a pragmatic and low-cost digital phenotyping approach that enables longitudinal tracking of common MS-related *patient-reported symptoms* in the patient’s own environment with minimal active patient engagement. Our approach harnesses passively collected sensor and behavior data from smartphones and fitness trackers and deploys machine learning models that achieve the highest prediction performance based on the most parsimonious data collection requirement. The key study finding is that, over 12 weeks (and 24 weeks in a subset), the best performing models achieved potentially clinically actionable accuracy (as well as F1-score, which summarizes positive predictive value and sensitivity) for predicting the short-term presence of depressive symptoms (every 2 weeks), high global MS neurological symptom burden, severe fatigue, and poor sleep quality (every 4 weeks) in pwMS, all significantly outperforming the baseline models. The best models for all four patient-reported symptoms included heart rate and steps as informative sensors.

For a secondary study goal, we report the marginal utility of behavioral features from the previous period (context features) in addition to behavioral features from the current period (action features) in helping the models contextualize an individual’s current behavior and in improving digital phenotyping of most common MS symptoms. For each patient-reported symptom, we performed pairwise comparisons of the six best models combining sensor or sensor plus EMA (comprising action-only versus action and context features, Figure 3) and operationally defined the “best” model as having the highest accuracy and F1-score while also requiring the least amount of sensor and/or EMA data. For predicting depressive symptoms, global MS neurological symptom burden, and sleep quality, the models containing action-only features were the “best” because the addition of context features did not improve the prediction of these patient-reported symptoms. In contrast, models containing action and context features improved the prediction of fatigue. Thus, behavioral features from longer periods that include context features (*i.e.,* the previous and current period) may have utility in the longitudinal symptom tracking of a smaller subset of common MS symptoms such as fatigue.

For another secondary study goal, we report the limited utility of incorporating minimal active patient input via EMA (*i.e.,* multiple choice response to two brief survey questions) into machine learning models in improving digital phenotyping of most common MS symptoms. For three of the four patient-reported symptoms (*i.e.,* global MS neurological symptom burden, fatigue, and sleep quality), the best models containing a combination of sensors plus average or pre-survey EMA did *not* significantly outperform the best models containing a combination of sensors without EMA. Thus, passively collected sensor data without any active patient engagement were sufficient to predict the severity of these patient-reported symptoms. For fatigue, this finding was particularly surprising given that one of the two administered EMA questions asked participants to rate the level of tired feeling. One possible explanation was that the EMA question might be too simplistic or insensitive to capture the complexity and dynamics of fatigue impact on the physical, cognitive and psychological function in pwMS [46, 47], though it was still unclear why a potentially relevant EMA did not improve the prediction for patient-reported fatigue severity at all. For depressive symptoms, the best models containing a combination of sensors plus average or pre-survey EMA *significantly outperformed* the best models containing a combination of sensors without EMA. While sensor data alone predicted depressive symptoms with reasonable accuracy (74.7%), the addition of pre-survey EMA yielded an 8.8% absolute increase in accuracy. This result was unsurprising given that the other EMA question asked participants to rate the level of depressed feeling. Notably, the best models containing pre-survey EMA were comparable to those containing average EMA, while pre-survey EMA (*i.e.,* the last EMA on the day before the patient-reported symptom survey) required substantially less active engagement by participants than average EMA (*i.e.,* three times daily). Overall, minimally active participant engagement may have some utility in the longitudinal symptom tracking of certain MS symptoms such as depressive symptoms.

Broadly, several aspects of our study differentiated from prior works, all with the goal of bringing digital phenotyping closer to clinical practice for pwMS. First, to demonstrate a basic feature of real-world applicability, our pragmatic study design leveraged each participant’s *own* digital device (*e.g.,* smartphone) to mitigate missing sensor data capture. In contrast, the earliest studies required a study-specific smartphone separate from participant’s own and increased participant burden [48]. Second, our approach passively harnessed data from a combination of multiple sensors in both smartphones and fitness tracker. Prior studies predicting MS outcomes based on passively sensed behavior largely relied on either a smartphone or fitness tracker (but not both) or a single sensor type [6, 7, 11, 13]. Third, our machine learning pipeline prioritized the *most parsimonious predictive models* containing the least amount of sensor and/or EMA data (*i.e.,* minimal or no active participant engagement) while still achieving clinically actionable accuracy and other prediction metrics. By comparison, most prior digital phenotyping efforts in MS prioritized performance without considering the amount of sensor data required for prediction and indeed often required active participant engagement, which would lead to lower adherence than passive sensing [9,48–55]. For instance, the study by Gashi *et al.* required participants to perform motor performance tests to classify fatigue levels in addition to passively sensed behavioral data [9]. Finally, our study outcomes as measured by validated survey instruments included a spectrum of common clinically relevant *patient-reported symptoms* that collectively reduce the quality of life in pwMS. On the other hand, standard clinical trial endpoints such as clinician-rated disability or functional testing scores [6, 8–9, 13–14, 56–64] as well as a single clinical outcome (at a time) [53, 65–68] in prior studies insufficiently captured the full real-world patient experience.

The current study also built on one of our own prior studies, which used passively sensed behavior changes during a state-mandated stay-at-home period (as compared to the pre-pandemic baseline) to predict depressive symptom, high global MS symptom burden, severe fatigue, and poor sleep quality in pwMS in a unique natural experiment in the setting of a global pandemic [19]. Specifically, we predicted the average value of patient-reported outcomes for each patient only once during a period (*i.e.,* the local COVID-19 stay-at-home period), whereas the current study made repeated clinical predictions (over consecutive 2 or 4-week periods) during 12 or 24-week study duration to emulate long-term symptom tracking in the real world. As methodological novelties, the current study further investigated the added utility of context behavioral features (from the previous periods) and two types of EMAs in improving digital phenotyping in MS.

Our digital phenotyping approach with *minimal or no active patient input* that reaches potentially clinically actionable prediction performance warrants additional investigations of its future clinical role in continuous tracking of *patient-reported symptoms* and in assisting comprehensive MS care in the real-world setting. Timely management of these common patient-reported symptoms could reduce delays in symptom management and greatly improve the quality of life for pwMS. Of clinical relevance, patient-reported symptoms assessed in this study are based on well-validated survey instruments that correlate with and complement clinician-rated outcomes. Practically, one can envision deploying continuous digital phenotyping to enable not only patient self-monitoring between routine clinic appointments but also crucial clinical triage for timely interventions (*e.g.,* medication initiation, counseling). Such approach may even be potentially useful in settings of limited healthcare access and resources though such clinical application would require dedicated testing.

Our study has at least two limitations. First, the study participant size, while larger than most previous digital phenotyping studies in MS, was still relatively modest. We made predictions for over 700 samples for depressive symptoms (in 2-week periods) and over 300 samples for global MS neurological symptom burden, fatigue and sleep quality (in 4-week periods) across 104 participants with MS. Notably, our well-characterized cohort also contrasts with larger studies where the diagnosis and/or patient-reported outcomes could not be independently verified [10, 54]. Crucially, we mitigated model over-fitting using leave-5-participants-out-cross-validation such that the participants used for training and testing were different in each fold. The consistently robust model performance across all five folds and for all four common MS patient-reported symptoms were reassuring. Second, we recruited study participants from a single clinic-based cohort, representative of its local MS patient population. Future validation in external cohorts with more racial and ethnically diverse patient populations would improve the generalizability of the approach.

In summary, our digital phenotyping approach using passively sensed data from their own smartphones and wearable fitness trackers could aid patients with real-world, continuous, self-monitoring of common symptoms in their native environment. It may also assist clinicians with better triage of patient needs for timely intervention in MS and potentially other chronic neurological disorders.

## Data Availability

All data produced in the present study are available upon reasonable request to the authors.

## AUTHOR CONTRIBUTIONS

Prerna Chikersal designed and conceptualized study; analyzed data; interpreted data; drafted and revised the manuscript for intellectual content. Shruthi Venkatesh and Elizabeth Walker played a major role in the data acquisition. Anind Dey and Mayank Goel designed and conceptualized study; interpreted the data; drafted and revised manuscript for intellectual content. Zongqi Xia designed and conceptualized study; major role in the data acquisition; interpreted data; drafted and revised the manuscript for intellectual content.

## ACKNOWLEDGEMENTS

We would like to thank our undergraduate research assistants: Man Jun (John) Han, Dong Yun Lee, Kasey Park, Phoebe Soong, and Christine Wu, for helping us monitor participant compliance throughout the data collection process. We would also like to thank Yiyi Ren for helping develop the app used for data collection. We would also like to thank the research participants and their treating clinicians.

The study was funded by the Department of Defense (CDMRP MS190178). In addition, the study was partially supported by grants from the National Institute of Health (NINDS R01NS098023 and NINDS R01NS124882).

## CONFLICTS OF INTEREST

None declared

## ABBREVIATIONS

MS: Multiple Sclerosis
EMA: Ecological Momentary Assessments
pwMS: People with Multiple Sclerosis
PHQ-9: Patient Health Questionnaire-9
MSRS-R: Multiple Sclerosis Rating Scale Revised
MFIS-5: Modified Fatigue Impact Scale-5
PSQI: Pittsburgh Sleep Quality Index
ML: Machine Learning
HR: Heart rate

## Notes

### Competing Interest Statement

The authors have declared no competing interest.

### Funding Statement

This study was partially funded by a Department of Defense grant and the National Institute of Health grants.

### Author Declarations

Ethics committee/IRB of the University of Pittsburgh gave ethical approval for this work. Ethics committee/IRB of Carnegie Mellon University gave ethical approval for this work.

### Summary of Updates

Added funding acknowledgments. Everything else is the same. Here is the funding information that is added: The study was funded by the Department of Defense (CDMRP MS190178). In addition, the study was partially supported by grants from the National Institute of Health (NINDS R01NS098023 and NINDS R01NS124882).

