## Supplementary Material for "Longitudinal Digital Phenotyping of Multiple Sclerosis Severity Using Passively Sensed Behaviors and Ecological Momentary Assessments"

#### A. Supplementary Methods

##### A.1. Details of Feature Extraction and Engineering

*Calls Features:* We used the call logs from the smartphone to calculate the following Calls features: *number and duration of all incoming, outgoing, and missed calls, number of correspondents overall*.

*Heart Rate Features:* We used the Fitbit Application Programming Interface (API) to calculate the following Heart Rate features: *mean heart rate, total time spent in the "fat burn" heart rate zone, total time spent in the "cardio" heart rate zone, total time spent in the "peak" heart rate zone, and total time spent below any heart rate zones indicating exercise (i.e., the out of the range zone).* The fat burn, cardio, and peak heart rate zones were calculated by Fitbit for each person separately.

*Location Features:* The Location 'virtual' sensor of the smartphone provided the best estimate of location based on available Global Positioning System (GPS), WiFi and cellular tower signals. We extracted the following Location features: *location variance* (sum of the variance in latitude and longitude coordinates), *log of location variance*, and *total distance traveled*. We then derived the following location-based features.

First, we used the Lomb-Scargle method [69] to calculate *Circadian movement* [45], which encoded the extent to which a person's location patterns followed a 24-hour circadian cycle.

Next, we labeled location samples as "static" or "moving" and clustered the "static" samples as described by Chikersal et. al. [29]. We then extracted several significant places, radius of gyration [35], percentage of time spent at top *(most frequented) places, percentage of time spent moving, and percentage of time spent in insignificant or rarely visited locations*. We further calculated the *average and standard deviation of length of stay at significant places* as well as *location entropy* and *normalized location entropy across significant places (*using previously described method [45]). Higher location entropy occurred when time is spent evenly across significant places.

Finally, we quantified features relevant to home and work locations. For operational reason, we assumed the place most visited by the participant at late night (between 00:00 hours to 06:00 hours) to be the *home location* and the place most visited by the participant during the afternoon (between 12:00 hours to 18:00 hours) to be the *work location*. To compute home location, we clustered the location coordinates from all nights and assumed the center of the most frequented cluster to be the participant's home location center. To compute work location, we clustered the location coordinates from all afternoons and assumed the center of the most frequented cluster to be the participant's work location center. As quality control, we used the self-reported home addresses of the participant to verify their home location, and switch home and work locations if the computed home location was found to not match with their self-reported home addresses. We calculated the *time spent at home, assuming home to be within 100 meters of the home location center* based on the default geofencing radius used by automation systems such as HomeKit (or Apple Home) and HomeAssistant https://www.home-assistant.io/.

*Screen Features:* We used the smartphone screen status sensor, which recorded screen status (on, off, lock, unlock) over time, to extract the following smartphone usage features: *total number of unlocks, mean number of unlocks per minute, total time spent interacting with the phone, and the median length of bouts (or continuous periods of time) during which the participant was interacting with the phone and when the screen was unlocked*. We defined that a participant was "interacting" with the smartphone during the interval between "unlocked" (on) and "locked" (off) screen status.

*Sleep Features:* We used the Fitbit API to calculate sleep features based on the daily sleep summaries and minute-to-minute sleep inferences (asleep, restless, awake, unknown). Sleep duration as captured by Fitbit is accurate ± 45 minutes [70-72]. First, we used each participant's entire sleep data to calculate the *total minutes asleep, total time in bed, and total sleep records*. Next, we excluded short naps and used the sleep data for the longest sleep record or "main" sleep period to calculate the *total time spent sleeping during main sleep, total time spent in bed during main sleep, sleep efficiency during main sleep,* which was calculated as (time asleep / [total time in bed - time to fall asleep])*, total time spent restless during main sleep, total number of times restless during main sleep, start time of main sleep in hours from midnight, and end time of main sleep in hours from midnight*.

*Steps Features:* We used the Fitbit API to calculate the steps features based on the step counts over time: *total number of steps, the number of minutes labeled as "sedentary" by Fitbit, the number of minutes labeled as "lightly active" by Fitbit, the number of minutes labeled as "fairly active" by Fitbit, and the number of minutes labeled as "very active" by Fitbit*.

#### A.2. Handing Missing Data

Missing sensor data could occasionally occur due to technical issues (*e.g.*, non-functioning phone / mobile application / server, faulty or delayed data transfer) or compliance issues (*e.g.*, participant not carrying the smartphone or wearing the FitBit). A missing feature during a time slice for many participants could indicate technical issues such as non-functioning server. A participant with many missing features could indicate technical issues such as the non-functioning phone or mobile application. A more likely reason for missing sensor data is the absence of activity. For example, if a participant made and received zero calls during a period, there would be no calls data. Thus, we encoded missing data into features since we could not differentiate whether such data were not collected or did not exist due to the absence of activity.

To empirically determine the thresholds, we plotted the number of participants and features remaining for various thresholds and noted the largest differential in curves (not shown). We then excluded all features in a time slice with missing values in more than 14 participants and likewise excluded participants missing more than 20% of all features. For each feature, we calculated the minimum feature value and imputed missing features as that value minus 1. Since we handled missing data independently across feature time slices, the number of participants and features differed across sensors and EMAs in each feature set.

### B. Supplementary Results

#### B.1. Results using the six 1-sensor models, the average EMA model, and the pre-survey EMA model

**Table S1**. F1-score (**accuracy in %**, n=sample size) of each of the 1-sensor models, the average EMA model, and the pre-survey EMA model.

| Sensors | Depression | | Global MS Symptom Burden | | Fatigue | | Sleep Quality | |
|---|---|---|---|---|---|---|---|---|
| | Action Only | Action + Context | Action Only | Action + Context | Action Only | Action + Context | Action Only | Action + Context |
| Calls | 0.58 (**41.2%**, n=729) | 0.58 (**41.2%**, n=729) | 0.66 (**49.1%**, n=338) | 0.66 (**49.1%**, n=338) | 0.58 (**57.5%**, n=339) | 0.59 (**58.1%**, n=339) | 0.60 (**43.0%**, n=321) | 0.60 (**43.0%**, n=321) |
| Heart Rate | 0.64 (**71.8%**, n=716) | 0.64 (**70.7%**, n=716) | 0.71 (**70.7%**, n=328) | 0.71 (**71.4%**, n=328) | 0.63 (**63.3%**, n=330) | 0.69 (**70.9%**, n=330) | 0.62 (**65.9%**, n=311) | 0.64 (**68.2%**, n=311) |
| Location | 0.57 (**67.2%**, n=740) | 0.56 (**65.9%**, n=740) | 0.66 (**68.4%**, n=342) | 0.63 (**65.2%**, n=342) | 0.65 (**48.7%**, n=343) | 0.65 (**48.7%**, n=343) | 0.56 (**63.1%**, n=322) | 0.51 (**59.9%**, n=322) |
| Screen | 0.57 (**40.2%**, n=757) | 0.57 (**40.2%**, n=757) | 0.65 (**48.3%**, n=348) | 0.65 (**48.3%**, n=348) | 0.63 (**65.3%**, n=349) | 0.59 (**61.6%**, n=349) | 0.58 (**65.6%**, n=328) | 0.58 (**64.9%**, n=328) |
| Sleep | 0.60 (**68.1%**, n=749) | 0.59 (**67.7%**, n=749) | 0.67 (**68.9%**, n=347) | 0.69 (**69.7%**, n=347) | 0.62 (**59.9%**, n=349) | 0.60 (**62.5%**, n=349) | 0.60 (**66.9%**, n=327) | 0.60 (**67.6%**, n=327) |
| Steps | 0.58 (**65.8%**, n=762) | 0.55 (**63.5%**, n=762) | 0.64 (**65.2%**, n=350) | 0.68 (**68.8%**, n=350) | 0.62 (**68.1%**, n=351) | 0.71 (**71.0%**, n=351) | 0.60 (**64.5%**, n=330) | 0.61 (**66.4%**, n=330) |
| Average EMA | 0.74 (**78.4%**, n=756) | 0.75 (**78.7%**, n=756) | 0.71 (**71.3%**, n=348) | 0.74 (**75.9%**, n=348) | 0.74 (**72.5%**, n=349) | 0.76 (**77.4%**, n=349) | 0.67 (**74.7%**, n=328) | 0.65 (**67.1%**, n=328) |
| Pre-Survey EMA | 0.75 (**78.6%**, n=756) | 0.75 (**78.8%**, n=756) | 0.67 (**65.8%**, n=348) | 0.70 (**69.8%**, n=348) | 0.75 (**72.2%**, n=349) | 0.74 (**75.1%**, n=349) | 0.65 (**66.5%**, n=328) | 0.65 (**66.5%**, n=328) |

Note:

1. "n=samples" is the number of samples in the dataset, indicating the number of periods for which we performed predictions across all participants.

##### B.2. Results obtained by combining ALL six sensors, ALL six sensors and average EMA, and all six sensors and pre-survey EMA

**Table S2.** F1-score (**accuracy in %**, n=sample size) of models combining all six sensors and/or each of the EMA type.

| Sensors | Depression | | Global MS Symptom Burden | | Fatigue | | Sleep Quality | |
|---|---|---|---|---|---|---|---|---|
| | Action Only | Action + Context | Action Only | Action + Context | Action Only | Action + Context | Action Only | Action + Context |
| All 6 Sensors | 0.68 (**74.7%**, n=668) | 0.66 (**72.2%**, n=668) | 0.72 (**70.7%**, n=311) | 0.72 (**72.0%**, n=311) | 0.64 (**60.4%**, n=313) | 0.71 (**69.7%**, n=313) | 0.56 (**58.2%**, n=297) | 0.64 (**68.7%**, n=297) |
| All 6 Sensors + Average EMA | 0.74 (**78.4%**, n=667) | 0.76 (**79.9%**, n=667) | 0.73 (**72.7%**, n=311) | 0.79 (**77.8%**, n=311) | 0.70 (**72.2%**, n=313) | 0.75 (**75.1%**, n=313) | 0.62 (**63.9%**, n=297) | 0.66 (**70.1%**, n=297) |
| All 6 Sensors + Pre-Survey EMA | 0.74 (**78.4%**, n=667) | 0.74 (**78.9%**, n=667) | 0.74 (**74.3%**, n=311) | 0.75 (**74.6%**, n=311) | 0.67 (**66.2%**, n=313) | 0.78 (**76.7%**, n=313) | 0.62 (**62.5%**, n=297) | 0.64 (**69.0%**, n=297) |

Note:

1. "n=samples" is the number of samples in the dataset, indicating the number of periods for which we performed predictions across all participants.
2. The model combinations included (i) the model that combines the predictions from all sensors, (ii) the model that combines the predictions from all sensors and the average EMA model, and (iii) the model that combines the predictions from all sensors and the pre-survey EMA model.

#### B.3. Best Sensor-EMA combinations that obtained the results reported in Figure 3

**Table S3.** The best combination of sensors and/or EMA for each model type.

| Model Types | Depression | Global MS Symptom Burden | Fatigue | Sleep Quality |
|---|---|---|---|---|
| **Action-Only & NoEMA** | ('calls', 'hr', 'loc', 'scr', 'slp', 'steps') | **('hr', 'loc', 'slp', 'steps')** [1] | ('calls', 'hr', 'scr', 'steps') | **('hr', 'loc', 'slp', 'steps')** [1] |
| **Action+Context & NoEMA** | ('calls', 'hr', 'loc', 'scr', 'slp') | ('hr', 'loc', 'slp') | **('hr', 'scr', 'steps')** [1] | ('calls', 'hr', 'slp', 'steps') |
| **Action-Only & AvgEMA** | ('calls', 'hr', 'loc', 'slp', 'avg_ema') | ('hr', 'loc', 'slp', 'steps', 'avg_ema') | ('hr', 'scr', 'steps', 'avg_ema') | ('hr', 'loc', 'scr', 'slp', 'avg_ema') |
| **Action+Context & AvgEMA** | ('calls', 'hr', 'loc', 'slp', 'avg_ema') | ('hr', 'loc', 'slp', 'steps', 'avg_ema') | ('hr', 'scr', 'slp', 'steps', 'avg_ema') | ('hr', 'loc', 'slp', 'steps', 'avg_ema') |
| **Action-Only & PresurveyEMA** | **('hr', 'steps', 'pre_survey_ema')** [1] | ('loc', 'slp', 'steps', 'pre_survey_ema') | ('hr', 'scr', 'steps', 'pre_survey_ema') | ('hr', 'loc', 'slp', 'steps') |
| **Action+Context & PresurveyEMA** | ('hr', 'loc', 'scr', 'pre_survey_ema') | ('hr', 'loc', 'scr', 'slp', 'pre_survey_ema') | ('calls', 'hr', 'scr', 'steps', 'pre_survey_ema') | ('hr', 'slp', 'pre_survey_ema') |

Note:

1. For each patient-reported symptom, we reported the model performance for each model type using the best combination of sensors or sensors and EMA models as shown in the main Figure 3.
2. *Abbreviations* - calls: Calls; hr: Heart Rate; loc: Location; scr: SmartPhone Screen Usage; slp: Sleep; steps: Steps; avg_EMA: Average EMA; pre_survey_EMA: Pre-survey EMA.

#### C. Data Availability

Code for analysis, summary and anonymous portion of the data for this study are available upon reasonable request to the corresponding author. Given the challenge of fully anonymizing sensor data, particularly the global positioning system tracking, sharing of full data set with external investigators will require IRB approval by the institution that governs the investigator making the request and institution-approved data transfer agreement.
